# Integrating Human Brain Proteomic Data with Genome-Wide Association Study Findings Identifies Novel Brain Proteins in Substance Use Traits

**DOI:** 10.1101/2022.02.02.22270270

**Authors:** Sylvanus Toikumo, Heng Xu, Rachel L. Kember, Henry R. Kranzler

## Abstract

**Background:** Despite the growing number of genetic risk loci identified for substance use traits (SUTs), the impact of these loci on protein abundance and their potential as therapeutic targets are unknown.

**Methods:** To address this, we performed a proteome-wide association study (PWAS) by integrating human brain proteomes from discovery (Banner; N = 152) and validation (ROSMAP; N = 376) datasets with genome-wide association study (GWAS) summary statistics for 4 SUTs. The sample comprised 4 GWAS of European-ancestry individuals for smoking initiation [Smk] (N = 1,232,091), alcohol use disorder [AUD] (N = 313,959), cannabis use disorder [CUD] (N = 384,032), and opioid use disorder [OUD] (N = 302,585). We conducted transcriptome-wide association studies (TWAS) with human brain transcriptomic data to examine the overlap of genetic effects at the proteomic and transcriptomic levels and tested significant genes for causality through Colocalization analysis.

**Results:** Twenty-seven genes (Smk=21, AUD=3, CUD=2, OUD=1) were significantly associated with cis-regulated brain protein abundance. There was evidence for causality in 6 genes (Smk: *NT5C2, GMPPB, NQO1, SRR*, and *ACTR1B;* AUD: *CTNND1*), which act by regulating brain protein abundance. Cis-regulated transcript levels for 8 genes (Smk=6, CUD=1, OUD=1) were associated with SUTs, indicating that genetic loci could confer risk for these SUTs by modulating both gene expression and proteomic abundance.

**Conclusions:** Functional studies of the high-confidence risk proteins (SRR for Smk and CTNND1 for AUD) identified here are needed to determine whether they are modifiable targets and useful in developing medications and biomarkers for these SUTs.

## Introduction

Substance use traits (SUTs), including smoking (Smk), alcohol use disorder (AUD), cannabis use disorder (CUD), and opioid use disorder (OUD0, are highly prevalent and leading causes of morbidity and mortality globally^1–3^. An estimated 40-60% of the risk of SUTs have been attributed to genetic factors^4–6^. Recent large-scale genome-wide association studies (GWAS) of SUTs have provided initial insights into their underlying biological systems^7–10^. Despite the growing success of GWAS in identifying associated single nucleotide polymorphisms (SNPs), the identified SNPs, many of which are intronic or intergenic^11^, exert only small effects, which suggests that their phenotypic effects are mediated by the regulation of gene transcription.

Recent advances aimed at understanding how SNPs influence gene transcription and contribute to disease pathogenesis have led to the development of analytic frameworks such as functional summary-based imputation (FUSION)^12^, S-PrediXcan^13^, summary data-based Mendelian randomization (SMR)^14^, and Coloc^15^. These frameworks utilize a transcriptome-wide association study (TWAS) approach, which integrates an external gene expression reference data and GWAS results to prioritize genes whose *cis*-regulated expression is associated with disease phenotypes.

To facilitate the identification of genes with *cis*-regulated expression profiles for SUTs, TWASs have been conducted for cigarette smoking^11^, cocaine dependence^11^, AUD^8^, and OUD^10^. Although these TWASs shed light on potential mechanisms through which genetic loci associated with SUTs exert their effects, the evidence they provide of expression quantitative trait loci (eQTL) effects are at the level of messenger RNA (mRNA), rather than protein abundance. Genetic variation can influence protein abundance by altering the rate and stability of gene expression^16^, though it remains to be determined whether the identified genetic loci exert their effects on SUTs by modulating protein abundance in the brain. The importance of this question lies in the fact that proteins, as the final products of gene expression, are the main functional components of cells and biological processes^17^, and comprise most drug targets and biomarkers^17,18^.

The key question addressed in the current study is whether loci identified through GWAS of SUTs contribute to their pathogenesis by modulating protein abundance. To answer this question, we applied an integrative proteome-wide association study (PWAS) approach that combines genetic data from four large GWASs of SUTs (including Smk, AUD, CUD, and OUD)^7–10^ with two independent human brain proteomic datasets (Banner^19^ and ROSMAP^20^) derived from brain dorsolateral prefrontal cortex (dPFC). To compare the effects of risk variants at both the proteomic and transcriptomic levels, we also performed TWAS using the CommonMind Consortium (CMC) dPFC^21^ and Genotype-Tissue Expression (GTEx) v7 frontal cortex^22^ datasets. Figure 1 provides an overview of the study.

**Figure 1:**
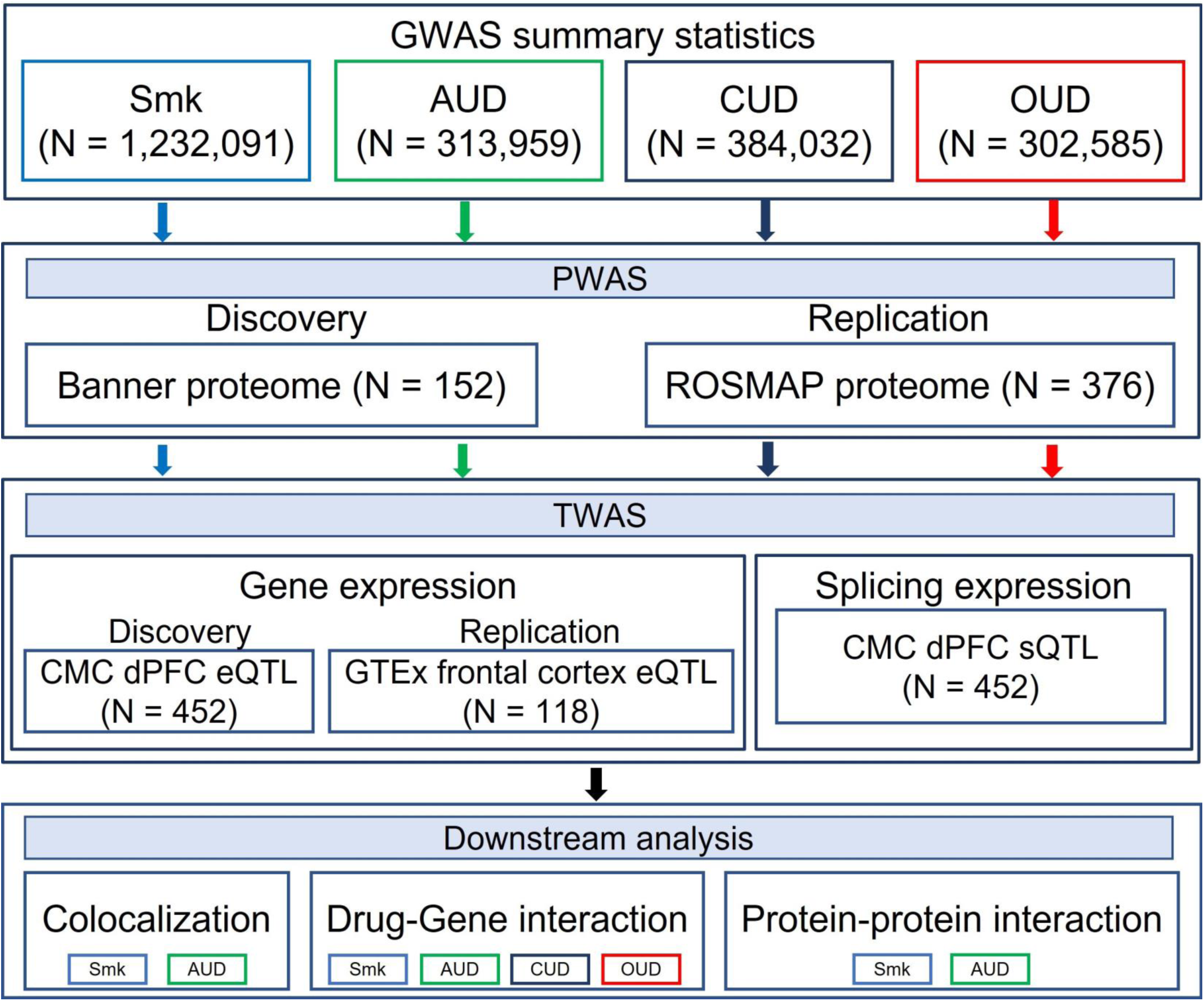
Overview of the study GWAS summary statistics included in the study were based on four substance use traits (SUT): smoking initiation (Smk), alcohol use disorder (AUD), cannabis use disorder (CUD) and opioid use disorder (OUD). For PWAS, human brain proteomes from Banner (discovery) and ROSMAP (replication) datasets were integrated with each set of GWAS summary statistics. TWAS based on brain eQTL datasets from discovery (CommonMind Consortium - CMC) and replication (Genotype-Tissue Expression - GTEx) datasets was conducted for each SUT. TWAS splicing expression analysis (CMC - sQTL) was also performed for all four traits. Colocalization analysis was based on nominally significant proteins and transcripts for Smk, and proteins for AUD. All significant proteins (after Bonferroni correction) were used as input for drug-gene interaction analysis for all four traits. Only proteome-wide significant proteins for Smk and AUD were included for Protein-protein interaction analysis.

## Methods

### Genome-wide association studies summary statistics

We selected the largest GWASs of SUTs that were available to us as of July 2021. The summary statistics were derived from 1,232,091 EUR for Smk^7^; 313,959 EUR for AUD^8^; 384,032 EUR for CUD^9^; and 302,585 EUR for OUD^10.^ We limited the GWASs to participants of European ancestry (EUR) to match the proteomic datasets. Study details including sample demographics and methods for phenotyping, data processing, and statistical analyses are provided in the original articles^7–10^ and summarized in Supplementary Table 1.

### Human brain pQTL data

We obtained human brain proteomic data from the study by Wingo et al.^23,24^, in which human protein abundance was quantified in the dorsolateral pre-frontal cortex (dPFC) of post-mortem brain tissues from 152 (Banner dataset)^19^ and 376 (ROSMAP dataset)^20^ EUR participants. By characterizing genetic control of the proteome in these human brain datasets, Wingo et al. identified 1,139 and 1,475 protein quantitative trait loci (pQTL) (hereafter referred to as protein weights) in the Banner and ROSMAP datasets, respectively. These protein weights were downloaded from http://doi.org/10.7303/syn23627957. Details on sample demographics, proteomic sequencing, quality control, and normalization can be accessed from Wingo et al^23,24^ and are summarized in Supplementary Table 1.

### Proteome-wide Association Analysis

To identify proteins whose genetically regulated expression is associated with SUTs, we performed PWAS analyses by integrating GWAS summary statistics of SUTs and pQTLs from a discovery (Banner) and validation (ROSMAP) datasets using the FUSION pipeline (http://gusevlab.org/projects/fusion/)^12^. For accuracy, FUSION employs 5-fold predictive models (top1, GBLUP, LASSO, Elastic Net, BSLMM) to compute the combined effect of SNPs on protein expression weights. The model with the largest cross-validation R^2^ was selected for downstream analyses. PWAS association statistics were Bonferroni corrected based on the number of proteins in the analysis (see the footnote in Table 1 and Supplementary Table 3).

**Table 1.** Results of the PWAS of smoking initiation.

| Gene | Chr | Banner |  | ROSMAP |  | Evidence for pQTL replication | Overlap with TWAS eQTL | Overlap with TWAS sQTL |
| --- | --- | --- | --- | --- | --- | --- | --- | --- |
|  |  | PWAS Z | PWAS p<sup>a</sup> | PWAS Z | PWAS p<sup>b</sup> |  |  |  |
| *NT5C2* | 10 | 7.29 | 3.09E-13 | 7.58 | 3.35E-14 | Yes | - | Yes |
| *ARPC1B* | 7 | 5.95 | 2.77E-09 | - | - | - | - | - |
| *HEBP1<sup>c</sup>* | 12 | -5.79 | 7.05E-09 | -0.68 | 4.94E-01 | No | - | - |
| *GMPPB* | 3 | 5.62 | 1.89E-08 | 5.01 | 5.36E-07 | Yes | - | - |
| *NQO1* | 16 | -5.32 | 1.02E-07 | -5.16 | 2.41E-07 | Yes | - | Yes |
| *SRR* | 17 | -5.04 | 4.73E-07 | -5.36 | 8.26E-08 | Yes | Yes | - |
| *RHOT2* | 16 | 4.95 | 7.50E-07 | 5.65 | 1.65E-08 | Yes | - | - |
| *ACTR1B* | 2 | -4.92 | 8.71E-07 | -4.77 | 1.82E-06 | Yes | - | - |
| *BTN3A3<sup>c</sup>* | 6 | -4.91 | 8.96E-07 | -3.24 | 1.19E-03 | No | - | - |
| *WIPI2<sup>c</sup>* | 7 | -4.23 | 2.34E-05 | -2.89 | 0.00382 | No | - | - |
| *BTN2A1* | 6 | 4.22 | 2.48E-05 | 4.55 | 5.26E-06 | Yes | - | - |
| *GFM1* | 3 | -4.20 | 2.66E-05 | - | - | - | - | - |
| *MAP1LC3A* | 20 | -4.10 | 4.12E-05 | - | - | - | - | - |
| *C10orf32* | 10 | - | - | -6.92 | 4.50E-12 | - | Yes | - |
| *TYW5* | 2 | - | - | 5.24 | 1.60E-07 | - | Yes | - |
| *MCTP1* | 5 | - | - | -4.85 | 1.21E-06 | - | - | Yes |
| *PLD1* | 3 | - | - | 4.66 | 3.13E-06 | - | - | - |
| *RFT1* | 3 | - | - | -4.66 | 3.19E-06 | - | - | - |
| *NAT6* | 3 | - | - | 4.64 | 3.55E-06 | - | Yes | - |
| *AAGAB* | 15 | - | - | 4.31 | 1.67E-05 | - | - | - |
| *PRKCD<sup>c</sup>* | 3 | 2.05 | 4.05E-02 | 4.25 | 2.14E-05 | No | - | - |

Chr - Chromosome; PWAS - proteome-wide association study; pQTL - protein quantitative trait loci; eQTL - expression quantitative trait loci; sQTL - splicing quantitative trait loci; ROSMAP - Religious Orders Study and Rush Memory and Aging Project
<sup>a</sup>Bonferroni correction p-value for Banner proteome-wide significant (PWS) genes was set at 4.36E-5
<sup>b</sup>Bonferroni correction p-value for ROSMAP PWS genes was set at 3.39E-5
<sup>c</sup>Genes that were PWS one human brain reference dataset and nominally significant (p < 0.05) in other brain proteome reference

### Human eQTL data

Human brain transcriptome data, used as expression reference panels, were obtained from the CMC^21^ and GTEx frontal cortex v7^12,22^. The CMC dataset consists of transcriptomic profiles for both gene-level (eQTL, n = 452) and intron-level (splicing – sQTL, n = 452) expression, which were generated from the dPFC^21^. CMC dPFC gene weights (eQTL and sQTL) and GTEx frontal cortex weights (eQTL, n = 136) were downloaded from the FUSION website (http://gusevlab.org/projects/fusion/)^12^.

To examine the association between the cis component of gene expression and SUTs, we performed a transcriptome-wide association analysis (TWAS) using the FUSION package^12^. TWAS was performed using gene and splicing expression profiles measured in the adult dPFC and gene expression profiles from the frontal cortex. For both PWAS and TWAS, we applied the default parameters recommended by FUSION. We also explored whether there was significant enrichment in the gene sets identified in the PWAS and TWAS. First, we identified genes that pass multiple correction testing in both PWAS and TWAS, defined as those which were significant in at least three analyses for PWAS-TWAS eQTL or two analyses for PWAS-TWAS sQTL. We then tested for significant overlap using a Binomial test (*p* < 0.05), as previously described^25^.

### Colocalization of PWAS and TWAS associations

To explore plausible causal relationships between GWAS variants and proteome- or transcriptome-wide associations, we performed colocalization analysis using the *coloc* R package (version 3.2-1)^15^ in FUSION^12^. We used the FUSION parameter (-coloc_P 0.05) to indicate the inclusion of nominally significant proteins/genes (at p < 0.05) and performed colocalization based on the GWAS and pQTL (ROSMAP and Banner)^23,24^, eQTL (CMC and GTEx)^21,22^ and sQTL (CMC)^21^ data. A posterior colocalization probability (PP) of 80% was used to denote evidence of a shared causal signal.

### Drug-gene interaction

We examined the proteins identified in the PWAS for known interactions with prescription drugs using the Drug Gene Interaction Database (DGIdb) v3.0 (https://www.dgidb.org)^26^. We categorized each identified prescription drug using the Anatomical Therapeutic Chemical (ATC) classifications obtained from the Kyoto Encyclopedia of Genes and Genomics Kyoto Encyclopedia of Genes and Genomics (KEGG: https://www.genome.jp/kegg/drug/).

### Protein-protein interaction

We used STRING database v11.0^27^ to assess whether PWAS genes were enriched for direct protein-protein interactions (PPIs). For these analyses, both discovery and replication proteome-wide significant (PWS) genes for Smk and AUD were used as input. STRING reports the confidence level for observed PPI using a scoring scheme (low confidence: < 0.4; medium: 0.4 – 0.7; high: > 0.7). We defined PWS genes within the observed PPI network as having the highest degree of network connections based on a STRING cut-off score > 0.4. We also used a whole genome reference model in STRING to determine whether the number of identified PPI were significantly enriched.

## Results

### PWAS identifies brain proteins for smoking initiation and other substance use traits

Using the FUSION pipeline to integrate pQTL and GWAS results to identify proteins whose abundance is correlated with the 4 SUTs^7–10^, in the discovery stage (using the Banner dataset) we identified 13 proteome-wide significant (PWS) genes for Smk (Table 1, Figure 2A) and 1 PWS gene each for AUD and CUD (Supplementary Table 2; Figure 1A). No gene was PWS for OUD in the Banner dataset (Figure 2A). Using the ROSMAP dataset for validation, we identified 15 PWS genes for Smk (Table 1, Figure 1B), 3 PWS genes for AUD (Supplementary Table 2, Figure 2B), and 1 PWS gene each for CUD and OUD (Supplementary Table 2, Figures 2B).

**Figure 2:**
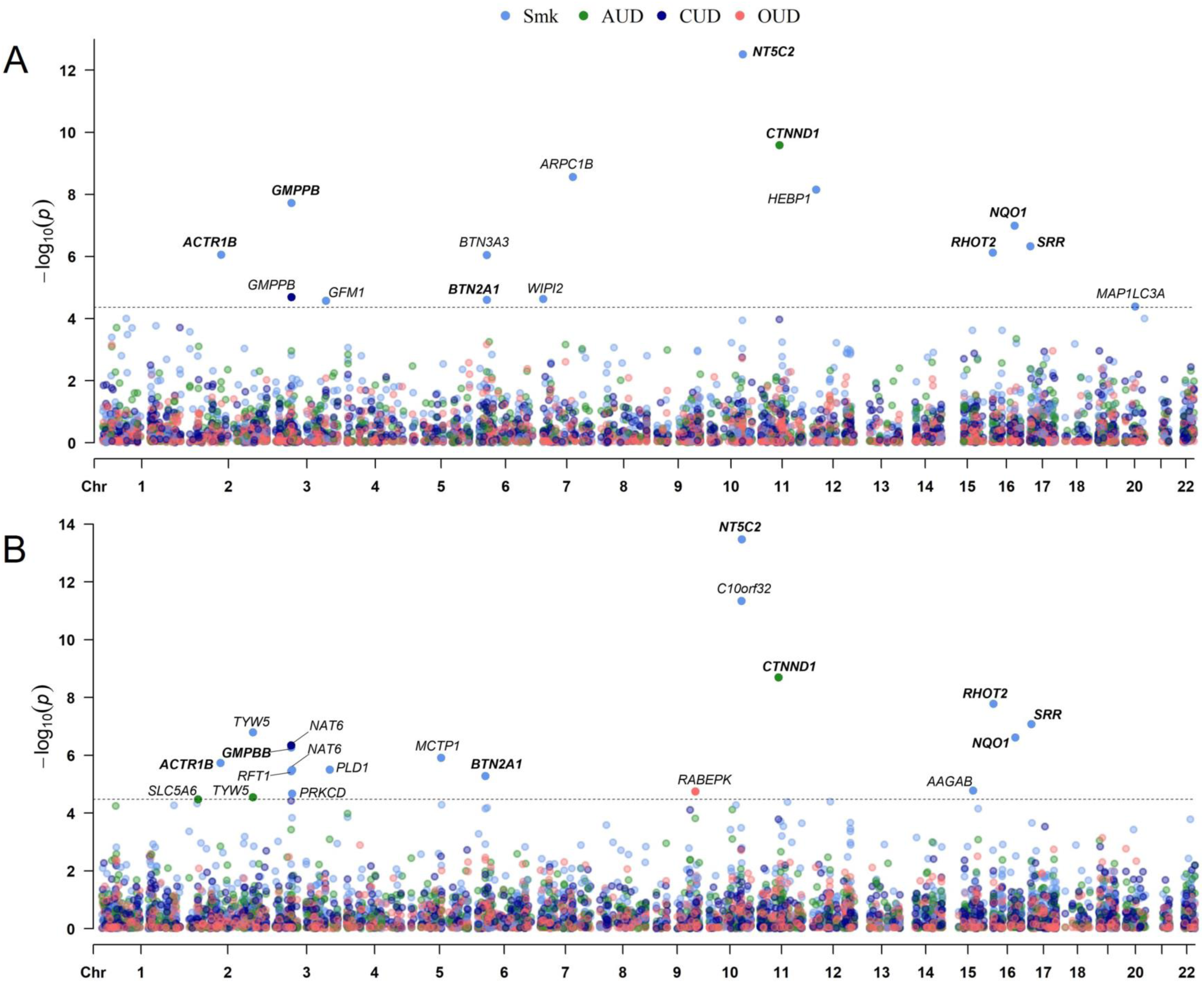
PWAS identified 27 genes and replicated 6 genes for substance use traits. (A) Manhattan plot for smoking initiation (Smk), alcohol use disorder (AUD), cannabis use disorder (CUD) and opioid use disorder (CUD) in the discovery proteome dataset. (B) Manhattan plot for Smk, AUD, CUD and CUD in the replication proteome dataset. Each dot on the x-axis denotes a gene and on the y-axis the strength of association (-log10 p-value). Proteome-wide significance level for discovery dataset; Bonferroni corrected p-value < 4.36 × 10^−5^ and replication; Bonferroni corrected p-value < 3.39 × 10^−5^. Replicated genes are in bold letters.

We next compared the PWS genes implicated in the discovery and replication stages for Smk, AUD, and CUD. Of the 13 high-confidence Smk PWS genes identified in the discovery Banner dataset (Table 1, Figure 2A), 7 were PWS (*NT5C2, GMPPB, NQO1, SRR, RHOT2, ACTR1B*, and *BTN2A1*) and 2 (*BTN3A3* and *WIPI2*) were nominally significant in the ROSMAP dataset (Table 1). The gene identified for AUD in the discovery dataset (*CTNND1*) was also PWS in the replication dataset (Supplementary Table 2) and the gene (*GMPPB*) that was PWS in the discovery cohort for CUD was near PWS (*p* = 3.75 × 10^−5^) in the replication dataset (Supplementary Table 2).

In addition to the replicated genes, 12 SUT genes were PWS in the replication stage only, including 8 genes for Smk (Table 1, Figure 2B), 2 genes for AUD and 1 gene each for CUD and OUD (Supplementary Table 2, Figures 2B). Of the 12 SUT risk genes identified in the replication stage, 1 was nominally significant for Smk (*PRKCD, p* = 4.05 × 10^−2^) (Table 1) and 1 for AUD (*SLC5A6, p* = 7.92 × 10^−4^) (Supplementary Table 2) in the discovery stage. In sum, by combining two independent human brain proteomic and SUT GWAS datasets, we identified 27 loci that could confer SUT risk through their effects on brain proteomic abundance.

### PWAS and TWAS overlap reveals high-confidence genes associated with SUTs

To identify SUT PWS genes with evidence of transcriptional regulation, we examined the extent of overlap at the protein and transcript levels. Specifically, we performed eQTL- and sQTL-based TWAS, followed by an analysis of the overlap between the transcriptome-wide significant (TWS) and PWS genes. For the discovery TWAS, we integrated SUT GWAS summary statistics^7–10^ and CMC dPFC eQTL datasets^21^. We detected 48 genes (38 for Smk, 6 for AUD, 2 for CUD, and 2 for OUD) whose cis-regulated expression was significantly associated with SUTs (Supplementary Tables, Supplementary Figures 1A - D) after Bonferroni correction. To validate these results, we also conducted TWAS using the GTEx frontal cortex eQTL dataset^22^, which identified 44 genes (36 for Smk, 4 for AUD, 2 for CUD, and 2 for OUD) with significant transcriptome-wide associations (Supplementary Tables 3, Supplementary Figures 2A – D).

Notably, 12 risk genes for Smk (*AS3MT, C10orf32, CPSF4, SFMBT1, SRR, ITIH4, TYW5, GPX1, CCDC88B, HYAL3, CNTROB*, and *NAT6*) from the discovery stage also showed TWS associations in the replication stage (Supplementary Table 3, Supplementary Figure 2B). For CUD, 2 TWS genes (*HYAL3* and *NAT6*) were replicated (Supplementary Table 3, Supplementary Figure 2B) while no TWS gene was replicated for AUD or OUD (Supplementary Table 3, Supplementary Figures 2C – D).

We next compared the replicated TWS eQTL genes with PWS genes (from both the discovery and replication stages) to ascertain whether there was significant overlap in SUT candidate risk genes. For Smk, 4 (*SRR, TYW5, C10orf32* and *NAT6*) of 21 PWS genes were confirmed by TWAS (Table 1, Supplementary Figure 3A), for which there was significant evidence of overlap with the binomial test (p = 3.58 × 10^−6^). One of the 2 PWS genes for CUD (Supplementary Table 2) – *NAT6* – was confirmed by TWAS (Supplementary Table 3). The binomial test was not run for CUD due to the small number of genes. No PWS gene was supported by TWAS for AUD (Supplementary Figure 3B) and OUD (Supplementary Tables 2 - 3).

At the level of splicing, we detected significant overlap between TWS sQTL genes and PWS genes for Smk (binomial test: p = 2.2 × 10^−16^; *NT5C2, NQO1*, and *MCTP1*) (Table 1, Supplementary Table 4). No overlapping genes were identified for AUD, CUD and OUD.

In sum, TWAS identified high-confidence genes with substantial evidence linking expression changes in *SRR, TYW5, C10orf32* and *NAT6*, and splicing of *NT5C2, NQO1*, and *MCTP1* to Smk risk.

### Colocalization of PWAS and TWAS genes

For the replicated genes in the proteomic (7 for Smk and 1 for AUD) and transcriptomic (7 for Smk) analyses, we explored whether there was evidence for a causal effect on SUTs. Colocalization analysis showed strong causal evidence for Smk in five PWS genes (*NT5C2, GMPPB, NQO1, SRR*, and *ACTR1B*) and six TWS eQTL genes (*AS3MT, TYW5, CCDC88B, CNTROB, SRR* and *C10orf32*) (coloc posterior probability (PP4) ≥ 80%; Supplementary Tables 5 – 9). We also found evidence of colocalization for the replicated AUD PWS gene (*CTNND1*) (PP4 ≥ 80%; Supplementary Table 10). These findings suggest that the same risk variants drive the associations between SUTs and both PWAS (for AUD and Smk) and TWAS eQTL (for Smk). None of the TWS sQTL genes were causal for Smk (Supplementary Table 9).

### Drug-gene and Protein-protein interaction

Because existing prescription medications can be repurposed to target encoded proteins, we queried all SUT risk genes (n = 27) that were detected by PWAS for interaction with prescription medications via DGIdb. We observed 33 interactions between 5 genes (*SRR, PRKCD, PLD1, NT5C2*, and *NQO1*) (Figure 3, Supplementary Table 11). *SRR*, which showed significant associations with Smk in PWAS and eQTL TWAS, was prioritized as a potential target of serine and pyridoxal phosphate in the antimycobacterials and vitamins drug classes, respectively. DGIdb also prioritized *NQO1*, a Smk risk gene in the PWAS and sQTL TWAS analysis, as a target of 15 drug interactions, which include analgesics (acetaminophen) and antiepileptics (cannabidiol).

**Figure 3:**
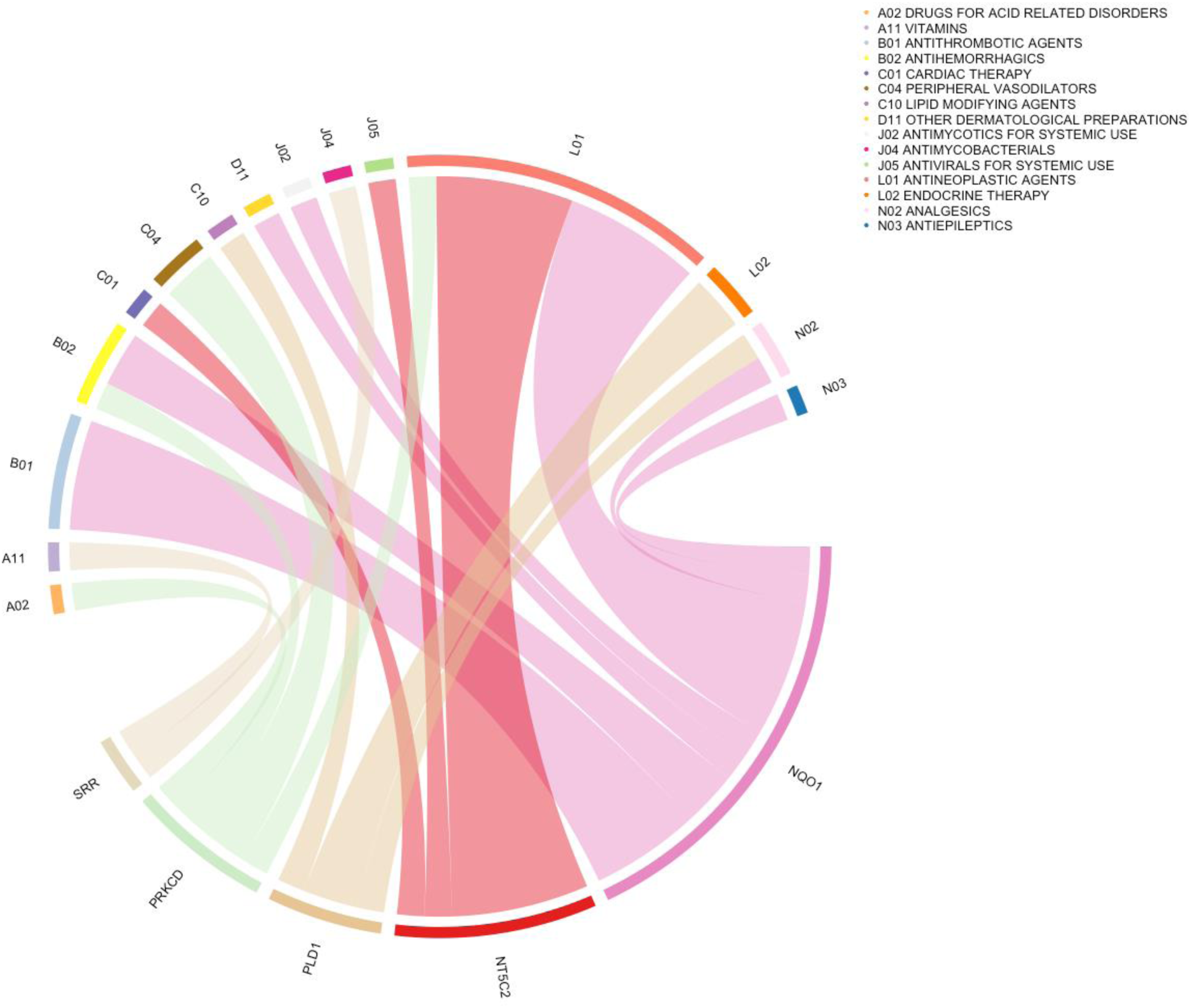
Drug-gene interaction prioritized 5 genes. Chord diagram of proteome-wide significant genes for SUTs and the Anatomical Therapeutic Chemical classification of drugs. Each gene is linked with drug classes and the width of each line is determined by the number of drugs in each class known to interact with each gene.

Direct protein-protein interaction was identified for two protein pairs (C10orf32 – NT5C2, interaction score = 0.567; MAP1LC3A – WIPI2, interaction score = 0.986) (Supplementary Table 12). However, these PPIs were not significantly enriched (*p* = 0.33), possibly due to the small number of proteins (N = 21) included in the PPI analysis or the limited proteomic reference information in the STRING database.

## Discussion

In this study, we sought to characterize the effect of genetic loci on the proteomic architecture of SUTs by performing PWASs that integrate human brain pQTL data^23,24^ with GWAS results for four SUTs^7–10^. We identified 27 SUT risk genes, of which 8 (*CTNND1* for AUD and *NT5C2, GMPPB, NQO1, SRR, RHOT2, ACTR1B* and *BTN2A1* for Smk) showed PWS associations in two independent brain proteomes. Notably, of the 8 replicated PWS genes, evidence for 6 (*CTNND1* for AUD and *NT5C2, GMPPB, NQO1, SRR*, and *ACTR1B* for Smk) was consistent with a causal effect based on Colocalization analysis. Thus, these genes could confer SUT risk by modulating protein abundance in the brain.

Three of the potentially causal genes identified at the protein level – *SRR, NT5C2*, and *NQO1* – showed significant associations with Smk at the transcript level. TWAS also identified 12 additional genes with evidence for eQTL replication, of which two showed PWS associations in one of the proteomic datasets for Smk (*C10orf32* and *NAT6*) and one for CUD (*NAT6 –* an eQTL in the CUD GWAS^9^) (Table 1). The greater number of PWS and TWS genes for Smk than other SUTs may reflect the larger sample size and genome-wide significant loci in the Smk discovery GWAS^7^. Although the number of TWS genes exceeds that for PWS genes identified for Smk, only about 30% of PWS genes overlap at the transcript level. This observation has been made in prior PWASs of psychiatric disorders^23,28^, and supports prior conclusions that mRNA transcript levels can explain between one-third and two-thirds of the variance in steady state protein abundance^29,30^. Moreover, mRNA and protein abundance levels are weakly correlated^31^ and have different genetic architectures^32^. Because gene expression is not a perfect proxy for protein expression^31^ studying brain proteins directly provides novel insights into the impact of genetic variation.

A proteomic effect for AUD was supported by the signal at *CTNND1*, a gene that harbors risk variants for anxiety disorder^33^, autism spectrum disorder^34^, and other neurodevelopment conditions^35^. The gene encodes a p120-catenin protein that is involved in regulating neuronal excitability and synaptic maturation^36^. Importantly, CTNND1 has been implicated as a risk protein in recent PWASs of depression^23,28^, a psychiatric disorder that shows high comorbidity and genetic overlap with AUD^37^. Our results, akin to other recent reports^23,28^, suggest that CTNND1 may confer shared risk on AUD and depression by affecting neuronal signaling and development.

Our Smk PWAS prioritized 5 causal genes, 3 of which (also supported by the TWAS) play roles in synaptic plasticity (*SRR*)^38,39^, neurodevelopment (*NT5C2*)^40,41^, and brain oxidative stress (*NQO1*)^42^. *SRR* encodes serine racemase, the enzyme that converts L- to D-serine, a co-activator of N-methyl-D-aspartate receptors (NMDAR), a key component in glutamatergic synaptic signaling in the brain^38,39^. *SRR* variants have been linked with prognosis in methamphetamine-induced psychosis^43^ and schizophrenia^44^. *SRR* deletion in mice reduces the cortical level of D-serine^45^, resulting in reduced NMDAR activation^46^, which has been associated with a reduced ability to extinguish conditioned responses to amphetamine^47^ and cocaine-associated stimuli^48^. Here, we report an association between smoking initiation and reduced SRR protein and transcript expression. We hypothesize that protein and expression changes in *SRR*, due to a shared risk variant, could mediate the adaptive processes involved in smoking initiation by altering NMDAR-dependent neurotransmission.

*NT5C2* encodes a phosphatase that interacts with adenosine monophosphate (AMP) to maintain cell proliferation and differentiation during neurodevelopment^40,41^. The gene regulates AMP-activated protein kinase (AMPK) signaling^41,49^ and harbors cis-eQTLs for Smk^50^ and schizophrenia^51^. Animal studies indicate that the AMPK signaling pathway is upregulated in mouse hippocampus following chronic nicotine exposure^52^. Our PWAS findings suggest that individuals who initiate smoking have a higher abundance in brain of the NT5C2 protein, which could negatively impact AMPK activity and, in turn, neuronal expression. Complementary to this notion, reduced *NT5C2* expression in fetal and adult dPFC, due to a common schizophrenia risk locus in *NT5C2*^51^, has been shown to disrupt AMPK signaling^41^. The underlying regulatory mechanism that mediates the effect of *NT5C2* on AMPK activation in the context of smoking behavior is not known and warrants further investigation.

As a member of the NADPH dehydrogenase (quinone) family, *NQO1* encodes a cytoplasmic 2-electron reductase that helps to regulate oxidative stress in brain by altering the level of reactive oxygen species in cells and by detoxifying carcinogens^53^. *NQO1* has been implicated in cigarette smoking^54,55^ and altered expression of *NQO1* in response to smoking has also been documented in animal^56^ and human^57^ studies. Exposure to cigarette smoke increases brain oxidative stress, thereby attenuating the brain defense mechanism in rats^58,59^ and mice^60^. In line with our findings that *NQO1* splicing and protein expression are associated with smoking initiation, a recent animal study revealed that cigarette smoke exposure is associated with upregulation of the antisense and mouse homolog of *NQO1* (*Nqo1-AS1*) in lung tissue of mice, resulting in attenuated oxidative stress *in vitro*^61^. Although requiring replication of these effects in brain, the findings suggest that *NQO1* expression changes can disrupt oxidative stress and contribute to the pathogenesis of smoking initiation.

Drug-gene interaction results prioritized pyridoxal phosphate (PLP), in the vitamins drug class, as a cofactor for SRR (the Smk risk protein), highlighting prior preclinical evidence that the human SRR is PLP-dependent^62–64^. As the metabolically active form of vitamin B6, PLP binds to SRR and stimulates NMDAR signaling, which is involved in brain metabolism and cellular antioxidant defense^62,63,65^. Tobacco smoke contains a substantial number of reactive oxygen species that could trigger oxidative stress in the brain^60^, blood-brain-barrier^66^ and periphery^67^, leading to vitamin B6 deficiency^67^. Of note, in humans, smoking reduced circulating plasma vitamin B6 and PLP levels^68,69^, with concentrations increasing significantly after a few days of smoking cessation^70^. This suggests that cigarette smoking could deplete circulating vitamin B6 and PLP levels by modifying the activity of the brain antioxidant defense triggered by PLP enzymatic interaction with SRR. Functional studies that investigate *SRR* as a druggable gene target for PLP enzyme activity following smoking exposure could provide a basis for the development of novel smoking-related treatments.

Our study should be interpreted in the context of limitations. First, the relatively small sample size from which the brain proteome reference dataset was derived, which contributed to an imbalance between pQTLs and eQTLs/sQTLs, limited our ability to capture the full spectrum of genetic effects on the proteome and transcriptome. This is reflected in the disparity between PWAS and TWAS results, in that larger samples in TWAS (CMC eQTL = 48/5419, GTEx eQTL = 44/3106, and CMC sQTL = 52/7771) provided higher statistical power for gene expression detection than for the PWAS (Banner = 15/1139; ROSMAP = 20/1475). Future PWAS of SUTs will require larger brain proteome datasets to permit better pQTL detection. Second, although we identified PWS and TWS genes and highlighted their plausible mechanistic pathways, we could not pinpoint the causal variants associated with these genes. Further efforts are needed to map the role of causal variants and functionally validate the described pathways in the context of SUTs. Thirdly, only participants of European descent were included in this study due to the lack of available data required to conduct the analyses in other population groups, thus limiting the generalizability of our findings. Future PWAS of SUTs should include samples from diverse populations but will depend on the availability of relevant reference data.

In conclusion, using PWAS, we identified 6 high-confidence genes that modulate brain protein abundance, thereby potentially altering biological pathways linked to the pathogenesis of SUTs. These genes are potentially modifiable targets for the development of medications and biomarkers for SUTs and thus warrant further investigation. These findings underscore the potential utility of the approach applied here to advance precision medicine efforts in diagnosing and treating SUTs.

## Data Availability

All data produced in the present study are available upon reasonable request to the authors

http://doi.org/10.7303/syn23627957

http://gusevlab.org/projects/fusion/

## Acknowledgments

This study was supported by the Veterans Integrated Service Network 4 Mental Illness Research, Education and Clinical Center and NIH grants DA046345, AA028292, and AA026364.

