## Supplementary Figures for "Integrating Human Brain Proteomic Data with Genome-Wide Association Study Findings Identifies Novel Brain Proteins in Substance Use Traits"

### Supplementary Figure 1: Results of TWAS CMC eQTL for substance use traits

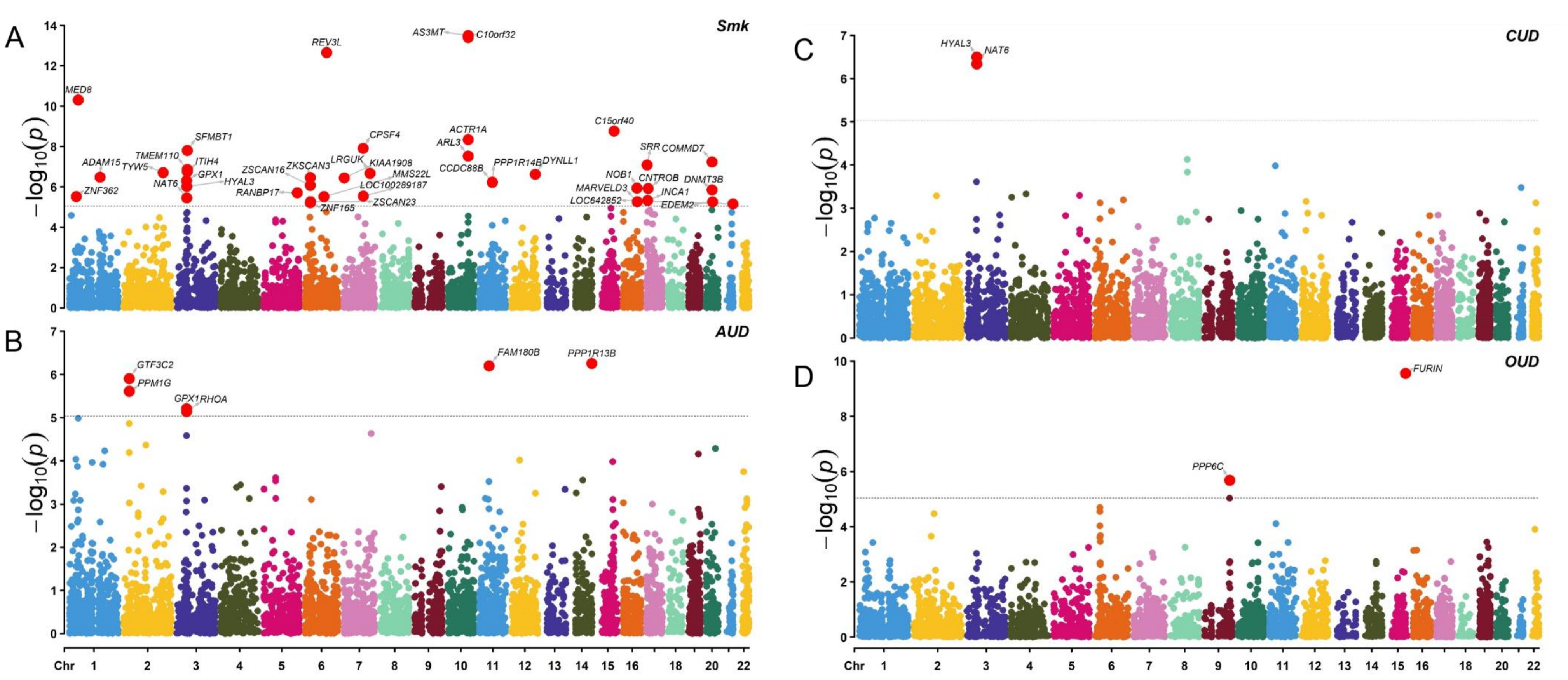


Manhattan plots for (A) smoking initiation (Smk); (B) alcohol use disorder (AUD); (C) cannabis use disorder (CUD); and (D) opioid use disorder (OUD). Transcriptome-wide association analyses were conducted using FUSION.

### Supplementary Figure 2: Results of TWAS GTEx eQTL for substance use traits

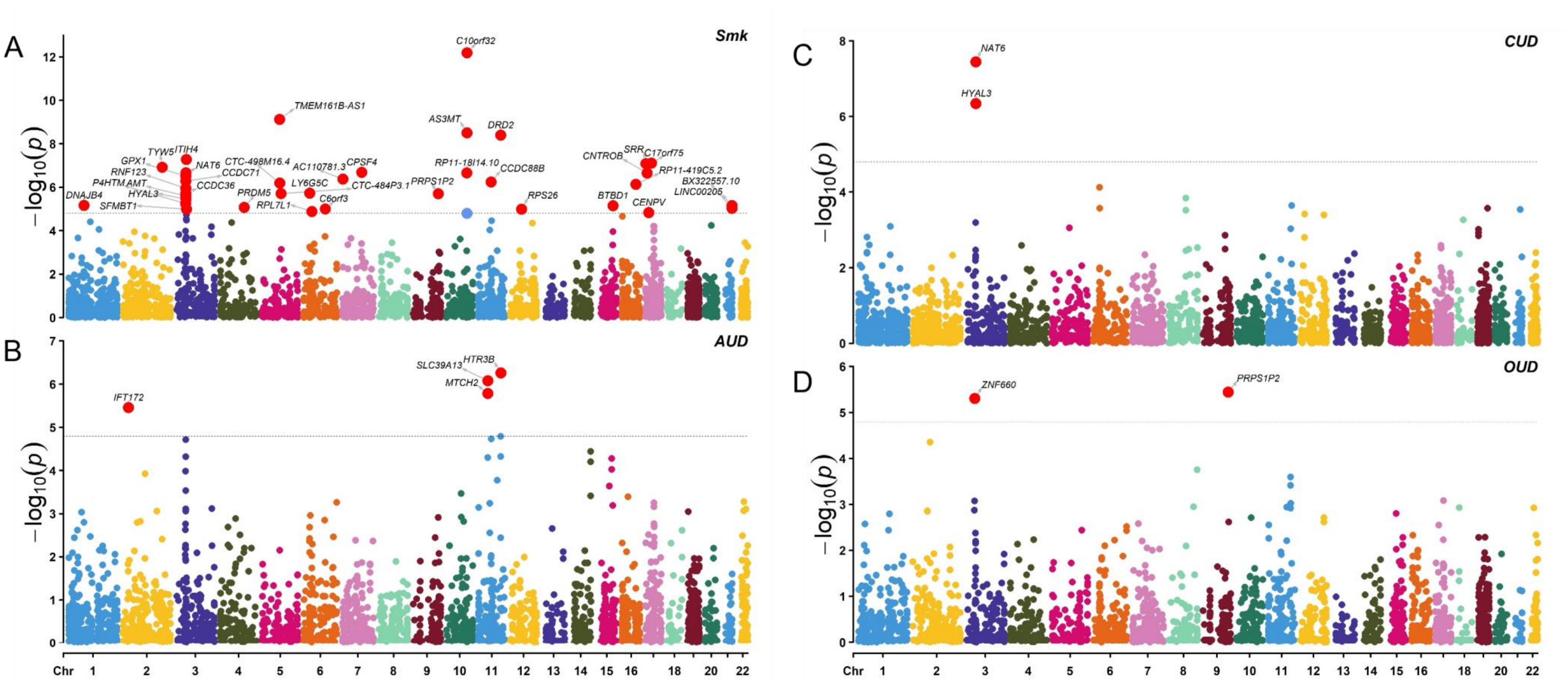


Manhattan plots for (A) smoking initiation (Smk); (B) alcohol use disorder (AUD); (C) cannabis use disorder (CUD); and (D) opioid use disorder (OUD). Transcriptome-wide association analyses were conducted using FUSION.

### Supplementary Figure 3: Gene overlaps for PWAS and TWAS eQTLs in substance use traits

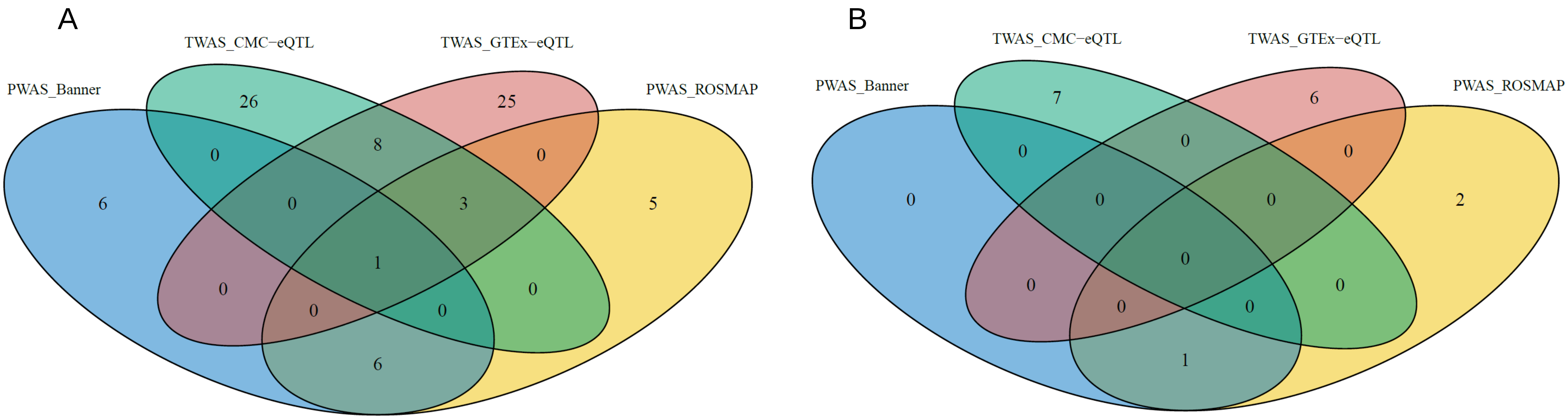
